# Head-to-head comparison between plasma p-tau217 and Flortaucipir-PET in amyloid-positive patients with cognitive impairment

**DOI:** 10.1101/2022.11.29.22282851

**Authors:** Nidhi S. Mundada, Julio C. Rojas, Lawren Vandevrede, Elisabeth H. Thijssen, Leonardo Iaccarino, Obiora C. Okoye, Ranjani Shankar, David N. Soleimani-Meigooni, Argentina L. Lago, Bruce L. Miller, Charlotte E. Teunissen, Hillary Heuer, Howie J. Rosen, Jeffrey L. Dage, William J. Jagust, Gil D. Rabinovici, Adam L. Boxer, Renaud La Joie

## Abstract

**Background and Objective:** Plasma phosphorylated tau (p-tau) has emerged as a promising biomarker for Alzheimer’s disease (AD). Studies have reported strong associations between p-tau and tau-PET that are mainly driven by differences between amyloid-positive and amyloid-negative patients. However, the relationship between p-tau and tau-PET is less characterized within cognitively impaired patients with a biomarker-supported diagnosis of AD. We conducted a head-to-head comparison between plasma p-tau217 and tau-PET in patients at the clinical stage of AD and further assessed their relationships with demographic, clinical, and biomarker variables.

**Methods:** We retrospectively included 87 amyloid-positive patients diagnosed with MCI or AD dementia who underwent structural MRI, amyloid-PET (^11^ C-PIB), tau-PET (^18^ F-flortaucipir, FTP), and blood draw assessments within one year (age=66±10, 48% female). Amyloid-PET was quantified in Centiloids (CL) while cortical tau-PET binding was measured using Standardized Uptake Value Ratios (SUVRs) referenced against inferior cerebellar cortex. Plasma p-tau217 concentrations were measured using an electrochemiluminescence-based assay on the Meso Scale Discovery platform. MRI-derived cortical volume was quantified with FreeSurfer. Mini-Mental State Examination (MMSE) scores were available at baseline (n=85) and follow-up visits (n=28; 1.5±0.7 years).

**Results:** Plasma p-tau217 and cortical FTP-SUVR were correlated (r=0.61, p<.001), especially in temporo-parietal and dorsolateral frontal cortices. Both higher p-tau217 and FTP-SUVR values were associated with younger age, female sex, and lower cortical volume, but not with APOE-ε4 carriership. PIB-PET centiloids were weakly correlated with FTP-SUVR (r=0.26, p=0.02), but not with p-tau217 (r=0.10, p=0.36). Regional PET-plasma associations varied with amyloid burden, with p-tau217 being more strongly associated with tau-PET in temporal cortex among patients with moderate amyloid-PET burden, and with tau-PET in primary cortices among patients with high amyloid-PET burden. Higher p-tau217 and FTP-SUVR values were independently associated with lower MMSE scores cross-sectionally, while only baseline FTP-SUVR predicted longitudinal MMSE decline when both biomarkers were included in the same model.

**Discussion:** Plasma p-tau217 and tau-PET are strongly correlated in amyloid-PET-positive patients with MCI and AD, and they exhibited comparable patterns of associations with demographic variables and with markers of downstream neurodegeneration.

## Introduction

Plasma measurements of phosphorylated tau (p-tau) have emerged as promising biomarkers for the detection of Alzheimer’s disease (AD) pathology in living patients. In the past three years, multiple studies have shown increased levels of plasma p-tau in patients with a clinical diagnosis of probable AD dementia compared to non-AD diagnoses or cognitively unimpaired individuals^1–6^. Cohorts with available autopsy information showed that *ante mortem* plasma p-tau concentrations were specifically increased in patients with neuropathologically-confirmed AD compared to patients with other etiologies^2–5,7–10^.

Blood-based biomarkers could constitute a less invasive, affordable, and scalable alternative to more established biomarkers derived from positron emission tomography (PET) or cerebrospinal fluid (CSF), with the potential to impact clinical diagnosis, large-scale research studies, and clinical trial design. Earlier studies showed strong associations between elevated p-tau concentrations and tau-PET as well as amyloid-PET positivity^1,5–7,9,11–14^. Beyond PET positivity, plasma p-tau levels are correlated with quantitative tau-PET measures; yet this association is partly driven by amyloid-negative cases having low values for both p-tau and tau-PET values^7,11^. Thus, it is not clear if p-tau and tau-PET are tightly coupled within the AD spectrum specifically, or if they merely both distinguish between AD and non-AD dementias. It is unknown whether plasma p-tau and tau-PET levels provide redundant or complementary information about disease severity, beyond their shared ability to detect the presence of AD pathophysiological processes.

We aimed to further characterize the relationship between tau-PET and plasma p-tau by focusing on these markers in amyloid-PET-positive patients at the early clinical stages of AD – similar to patients included in multiple clinical trials^15,16^. Based on the extensive literature showing that tau-PET consistently correlates with patient demographics (e.g. age^17–19^ and sex^20,21^) and downstream pathological markers (e.g. clinical deficits^22^, brain volume^23,24^) in amyloid-positive patients, we aimed to determine whether plasma p-tau showed similar patterns of associations. While multiple plasma p-tau biomarkers have been investigated (p-tau-181, 217, 231, based on the phosphorylation site), we focused on plasma p-tau217 because it has shown strongest associations with PET and neuropathological measures of AD^7,25^. We tested associations of plasma p-tau217 and tau-PET with patient demographics, amyloid-PET burden, APOE-ε4 carriership, as well as with two markers of putatively downstream pathophysiological processes: brain atrophy and cognitive impairment. Based on existing literature on tau-PET and biofluid biomarkers^5,26^, we hypothesized that: (i) Tau PET-plasma correlations would be relatively weak within amyloid-positive, cognitively impaired patients; and (ii) Tau-PET would exhibit stronger associations than plasma p-tau217 with downstream disease markers.

## Methods

### Patients

Patients were retrospectively selected from the University of California San Francisco (UCSF) Alzheimer Disease Research Center. Based on our specific goals, we selected patients who fulfilled the following criteria (Supplementary Figure 1): (1) available plasma sample with measurement of p-tau217; (2) clinical diagnosis of Mild Cognitive Impairment^27^ (Clinical Dementia Rating, i.e., CDR = 0.5) or AD dementia (CDR = 1 or higher) due to AD^28^ ; (3) available MRI and PET with both [^11^ C]-Pittsburgh compound B (PIB) for amyloid and [^18^ F]-flortaucipir (FTP) for tau within one year of plasma sample; (4) PIB-PET visually read as positive; and (5) no known genetic variants associated with autosomal dominant neurodegenerative diseases. These criteria resulted in a final sample of 87 patients; all were previously included in a larger study that included both AD and non-AD clinical diagnoses and biomarker profiles^7^.

Eighty-five patients had Mini-Mental State Exam (MMSE) scores available within 1 year of blood sample collection. Twenty-eight patients (33%) also had follow-up MMSE scores which were used to measure cognitive decline prospectively (116 observations from 85 patients: 57 patients with baseline MMSE only, 25 patients with baseline + 1 follow up, 3 patients with baseline + 2 follow ups; time between baseline and last MMSE = 1.5±0.7 years).

### Standard Protocol Approvals, Registrations, and Patient Consents

Patients provided written, informed consent at the time of recruitment. The study was approved by the institutional review boards at UCSF, University of California, Berkeley, and Lawrence Berkeley National Laboratory.

### Biomarker and Imaging Measurements

#### Imaging Acquisition

Patients underwent a 3T MRI at UCSF on either a 3T Siemens Tim Trio (n=23) or a 3T Siemens Prima Fit (n=64) scanner. T1-weighted magnetization-prepared rapid gradient-echo MRI sequences (sagittal slice orientation; 1 × 1 × 1 mm resolution; slices per slab = 160; matrix= 240 × 256; repetition time = 2.3 ms; inversion time = 900 ms; flip angle = 9° ; echo time = 2.98 ms for Trio and 1.9 for Prisma) were used for PET preprocessing and to extract cortical volume.

PET scanning was performed at the Lawrence Berkeley National Laboratory on a single Siemens Biograph PET/CT scanner. PIB and FTP were synthesized and radiolabeled locally. We analyzed PET data that were acquired from 50 to 70 minutes post-injection of ∼15 mCi of PIB (four, 5-minute frames) and 80 to 100 minutes post-injection of ∼10 mCi of FTP (four, 5-minute frames). A low-dose CT scan was acquired for attenuation correction prior to PET acquisition, and PET data were reconstructed using an ordered subset expectation maximization algorithm with weighted attenuation and scatter correction and a 4 mm Gaussian kernel applied during reconstruction (image resolution:6.5 × 6.5 × 7.25 mm estimated based on Hoffman phantom).

#### Imaging Processing

T1 MRIs were segmented and parcellated using FreeSurfer version 5.3 (surfer.nmr.mgh.harvard.edu). PET frames were realigned, averaged, and coregistered onto their corresponding MRI using Statistical Parametric Mapping 12 (Wellcome Department of Imaging Neuroscience, Institute of Neurology, London, UK). Standardized uptake value ratio (SUVR) images were created using tracer-specific reference regions: cerebellar gray matter for PIB-PET and inferior cerebellar gray matter for FTP-PET^29^.

To obtain a measure of global cortical Aβ and tau burden, we extracted a mean, cortical SUVR value for each tracer in native space using a weighted average of all FreeSurfer-derived cortical regions. PIB-PET SUVR values were converted to centiloids (CL) based on a previously validated equation,^30^ and scans were independently read as positive by expert readers blind to clinical or plasma biomarker information^31^.

Finally, we extracted cortical gray matter volume and total intracranial volume (TIV) for each patient using FreeSurfer. Adjusted cortical volume was calculated as 100 * (cortical gray matter volume / TIV).

#### Biomarker Measurements

Plasma p-tau217 concentrations were measured using electrochemiluminescence-based assays on the Meso Scale Discovery platform (MSD, Rockville, MD, USA). Biotinylated-IBA493 was used as a capture antibody and 4G10E2 as the detector antibody for the Eli Lilly p-tau217 assay^7^.

### Statistical Analysis

We calculated the Pearson correlation between mean cortical FTP-SUVR and p-tau217 concentration across patients. To determine if the correlation between cortical FTP-SUVR and p-tau217 was driven or modulated by patient demographic variables or cortical amyloid burden, we performed multiple linear regression analyses with cortical FTP-SUVR as the dependent variable, p-tau217 as an independent variable, and (in separate models, due to limited sample size) age, sex, APOE-ε4 carriership, or cortical PIB-CL value as a second independent variable. We also included the interaction between p-tau217 and the second independent variable. For each model, we then considered: (1) If p-tau217 remained a significant main effect in the presence of a covariate (e.g., age) and interaction; and (2) If the interaction between p-tau217 and this covariate (e.g., p-tau217*age) was a significant predictor of FTP-SUVR.

We explored the associations of each tau biomarker with demographic and biomarker variables: age and PIB-CL using bivariate correlations, sex using independent samples t-tests, and APOE-ε4 carriership (coded as non-carrier, heterozygote, and homozygote) using one-way analyses of variance (ANOVA). To assess whether these variables were more strongly associated with one tau biomarker than the other, we computed 95% confidence intervals of the difference for the corresponding effect sizes (e.g., r_age-PET_ versus r_age-plasma_) using bootstrapped resampling (N=1000 iterations), see Supplementary material for example R code.

We performed voxelwise analyses to characterize regional associations between p-tau217 and tau-PET. In addition, we looked at whether these regional association patterns varied with age, sex, APOE-ε4 carriership, or PIB-CL by adding an interaction term to our voxelwise models (e.g., dependent variable: FTP-SUVR in each voxel; independent variables: p-tau217, sex, p-tau217*sex). Voxelwise analyses were considered statistically significant using an uncorrected p<0.001 peak thresholds combined with a cluster size > 100 voxels. In addition, models were assessed using a more stringent threshold of family wise error (FWE) corrected p<0.05.

A key goal of this study was to describe and compare the association of plasma and PET tau biomarkers with indices of downstream pathophysiological processes. We considered measures of neurodegeneration (adjusted cortical volume) and cognitive impairment (MMSE score) as outcomes of interest. Using cross-sectional data, we ran multiple general linear models to determine whether tau-PET and p-tau217 provided redundant, or complementary information with respect to adjusted cortical volume and MMSE score. For each outcome variable (i.e., cortical volume or MMSE), we ran three models using (1) mean cortical FTP-SUVR; (2) p-tau217; or (3) both biomarkers as independent variables of interest. All models also controlled for age and sex. Model fitness was assessed using R^2^ (higher is better) and the Akaike Information Criterion (AIC, lower is better), which discourages overfitting by penalizing models with more independent variables.

Finally, we assessed the relationship between baseline tau biomarkers and subsequent change in MMSE scores using fixed-slope, random-intercept, linear mixed-effects models that included all available baseline and longitudinal MMSE data. All models included MMSE as the dependent variable, time from baseline (in years, non-centered) as a fixed covariate, and subject as a random effect. Similar to cross-sectional analyses, three alternative models were run, each with different combinations of added fixed effects: (1) mean cortical FTP-SUVR and FTP-SUVR*time interaction; (2) p-tau217 and p-tau217*time interaction; and (3) FTP-SUVR, p-tau217, and their respective interactions with time. Biomarker values were mean-centered; intercepts therefore represent predicted MMSE at baseline for average biomarker values. See Supplementary material for R code.

Statistical analyses were performed using R (version 4.1.1).

## Data Availability

Data that support the findings of this study are available upon request (memory.ucsf.edu/research-trials/professional/open-science). Voxelwise results are publicly available on neurovault (https://neurovault.org/collections/NLWHVBKP/).

## Results

Eighty-seven patients were included in the study and covered a large age range (49 – 95 years old, mean = 66.4, SD = 9.6), 48% female, and 95% White (Table 1). In the total sample, 53 (61%) patients had a clinical diagnosis of MCI and 44 (39%) of AD dementia; all patients were amyloid-PET positive per inclusion criteria. The average time between blood draw and FTP-PET was 69 (SD = 80) days, PIB-PET was 65 (SD = 72) days, and baseline MMSE score was 8 (SD = 35) days.

**Table 1.**
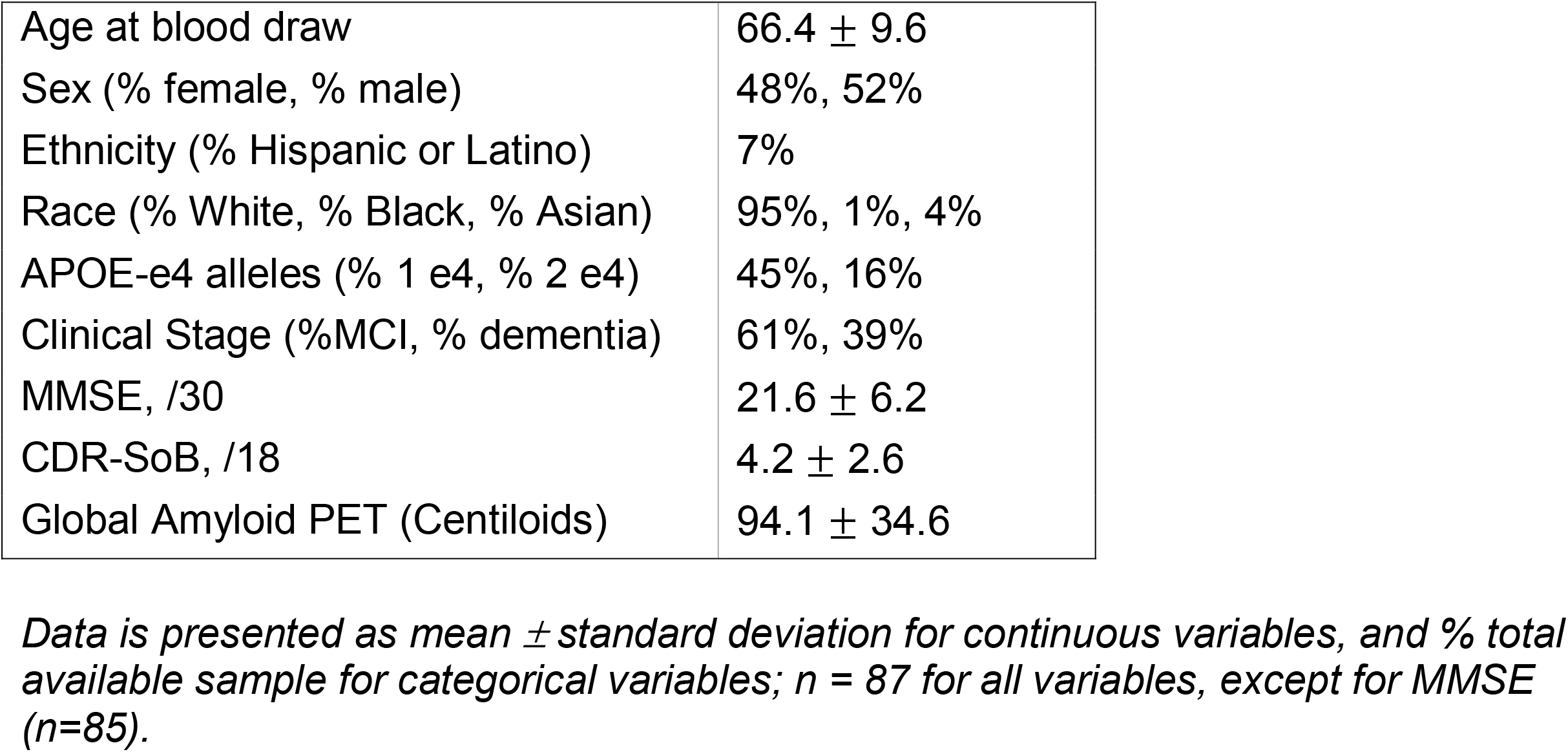
Sample characteristics (n=87)

### Association between tau biomarkers and demographics

In the whole group, cortical FTP-SUVR and p-tau217 concentration were correlated (r=0.61, p<0.001, 95% CI: 0.45-0.72). The residuals of this correlation were highly heteroscedastic, with greater variability at higher biomarker values (Figure 1); the right panel of figure 1 illustrates the variability of tau-PET scans in patients with a similar p-tau217 concentration value of ∼0.7 pg/mL, with various levels of overall binding and great heterogeneity in regional patterns. The correlation between FTP-PET and p-tau217 remained significant in the presence of added covariates for age, sex, APOE-ε4 carriership, and Centiloids (tested separately; see Methods). In addition, no interaction between p-tau217 and any of these covariates was a significant predictor of FTP-PET (p-values all >0.25). Cortical FTP-SUVR and p-tau217 were therefore closely related across patients, and this relationship was not explained or moderated by demographic variables or amyloid burden.

**Fig 1.**
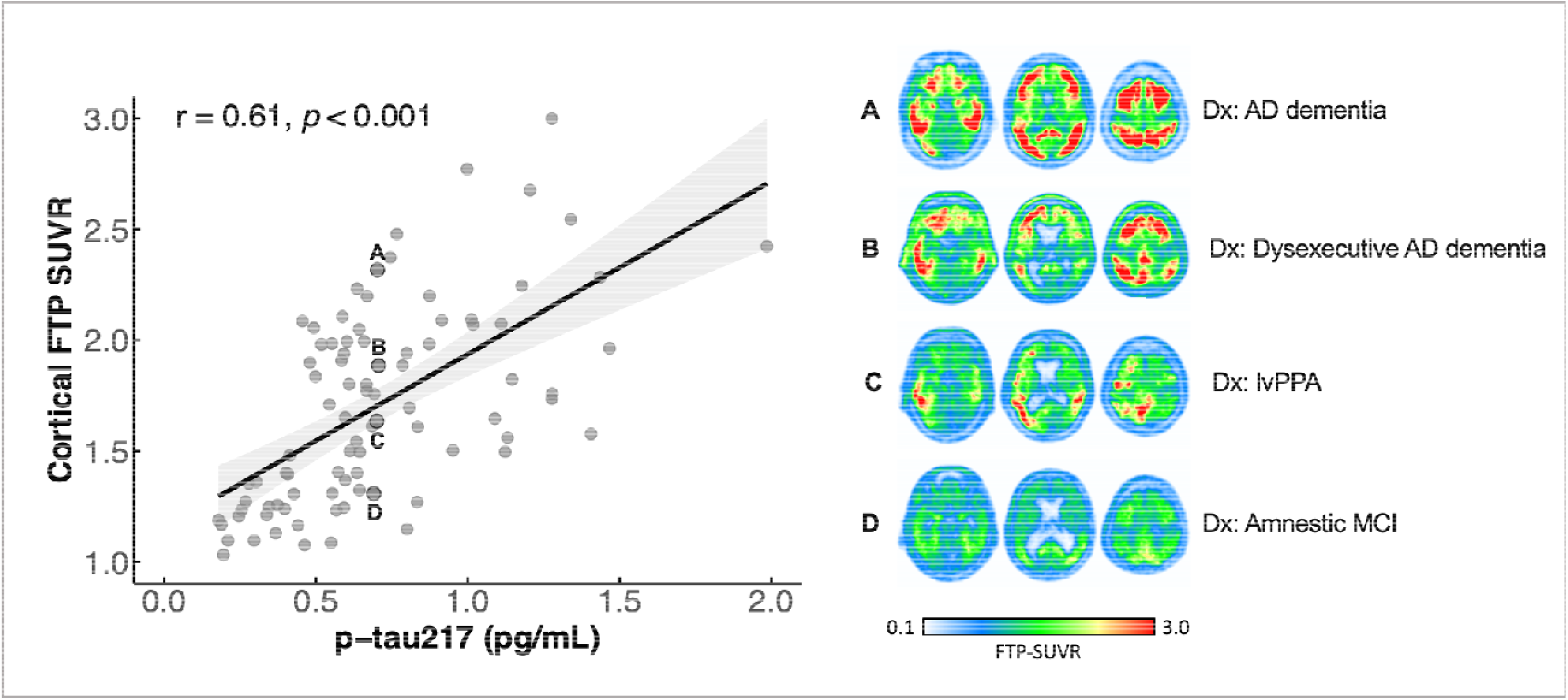
Correlation between p-tau217 and FTP-PET. Each scatterpoint shows data from one patient. The shaded area indicates 95% CI of the regression line. Data points labelled A, B, C, D correspond to patients with ∼0.7pg/mL p-tau217 concentration; their corresponding FTP-PET SUVR maps are shown on the right panel.

Next, we assessed if FTP-SUVR and p-tau217 showed similar patterns of associations with demographics. Both tau biomarkers were higher in younger patients (FTP-SUVR: r=-0.68, p-tau217: r=-0.44, both p’s<0.001; Figure 2A) and females (FTP-SUVR: d=0.78, p-tau217: d=0.53, both p’s<0.001; Figure 2B). In contrast, biomarker values were independent of APOE-ε4 carriership (d’s<0.22, p’s>0.32; Figure 2C). Lastly, amyloid-PET CLs were weakly correlated with FTP-SUVR (r=0.26, p=0.02), while there was no significant association between amyloid-PET CLs and p-tau217 (r=0.10, p=0.36; Figure 2D). Although associations tended to be stronger for PET compared to plasma, bootstrap-based comparisons of the associations were only significant for age (p=0.003), but not for sex (p=0.21), APOE-ε4 carriership (p=0.81), or Centiloids

**Fig 2.**
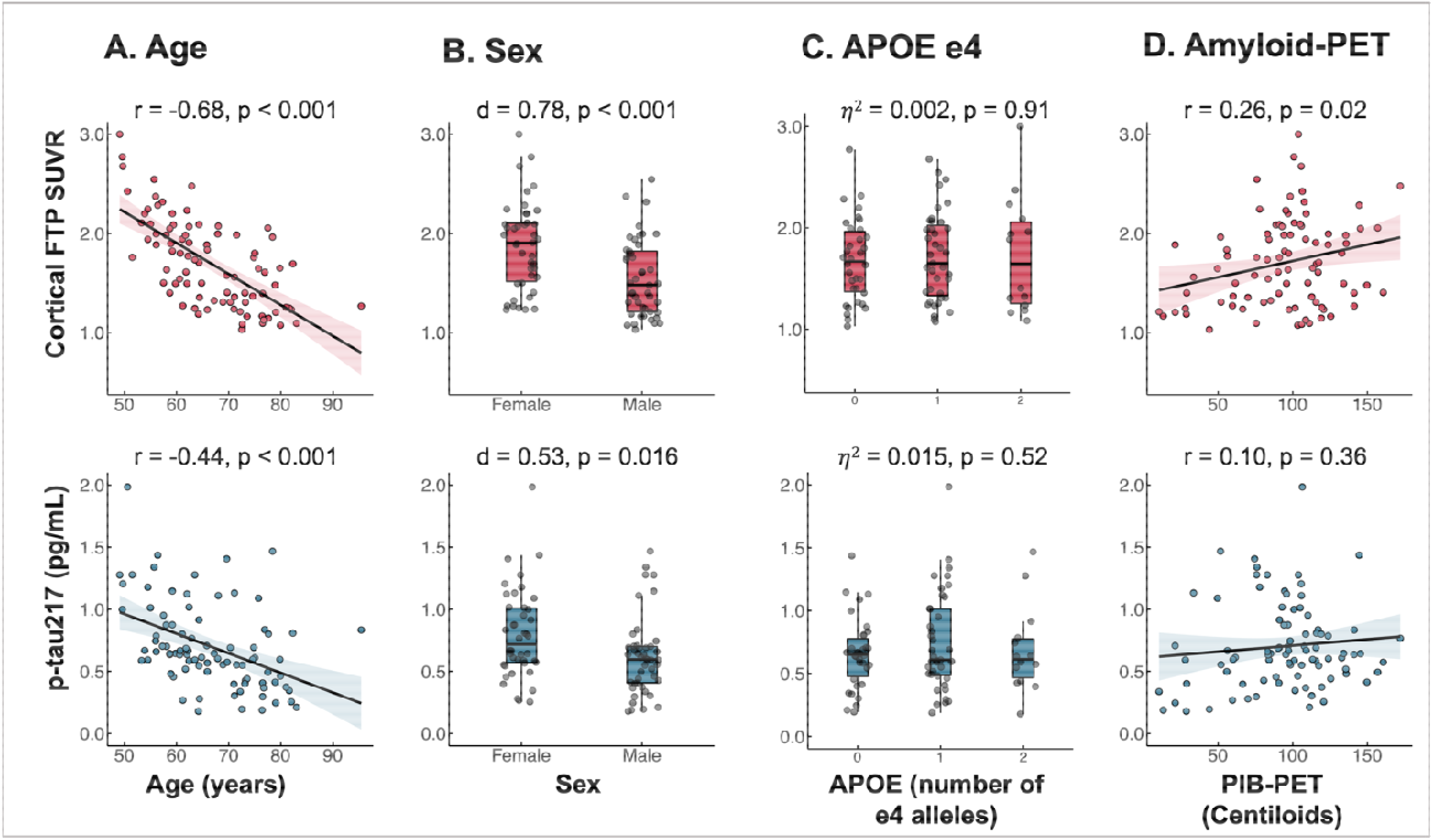
Association of tau biomarkers with age, sex, APOE4, and amyloid-PET levels. The relationship between cortical FTP-SUVR (top row) and p-tau217 (bottom row) is shown with respect to patient age (A), sex (B), APOE-ε4 (C), and cortical amyloid-PET uptake (D). Each scatterpoint shows data from one patient. r = Pearson’s correlation coefficient; d = Cohen’s d effect size; 2 = eta square. In panels A & D, the shaded area indicates 95% CI of the regression line.

(p=0.09); see Supplementary Table 1 for full results and Supplementary Methods for corresponding R code. Patterns of associations remained unchanged after log-transformation of biomarker values or using rank-based statistics (Supplementary Table 2).

**Table 2.** Cross-sectional associations between tau biomarkers and downstream measures of neurodegeneration and cognitive impairment.

|  | Model 1: PET only |  |  | Model 2: plasma only |  |  | Model 3: both |  |  |
| --- | --- | --- | --- | --- | --- | --- | --- | --- | --- |
| **A. Adj. Cort. Vol.** | $\beta$ | 95% CI of $\beta$ | p | $\beta$ | 95% CI of $\beta$ | p | $\beta$ | 95% CI of $\beta$ | p |
| Age | -0.07 | [-0.13, -0.02] | **0.008** | -0.04 | [-0.09, 0.002] | 0.063 | -0.08 | [-0.13, -0.02] | **0.007** |
| Sex | 1.29 | [0.47, 2.12] | **0.002** | 1.08 | [0.27, 1.89] | **0.010** | 1.31 | [0.49, 2.14] | **0.002** |
| FTP SUVR | -1.75 | [-2.99, -0.52] | **0.006** |  | -- |  | -1.46 | [-2.84, -0.09] | **0.037** |
| p-tau217 |  | -- |  | -1.33 | [-2.62, -0.05] | **0.043** | -0.68 | [-2.08, 0.72] | 0.336 |
| R<sup>2</sup> | 0.165 |  |  | 0.129 |  |  | 0.174 |  |  |
| AIC | 354 |  |  | 358 |  |  | 355 |  |  |
| **B. MMSE** | $\beta$ | 95% CI of $\beta$ | p | $\beta$ | 95% CI of $\beta$ | p | $\beta$ | 95% CI of $\beta$ | p |
| Age | -0.08 | [-0.24, 0.07] | 0.302 | 0.06 | [-0.07, 0.19] | 0.358 | -0.10 | [-0.25, 0.05] | 0.205 |
| Sex | 2.68 | [0.28, 5.07] | **0.029** | 1.75 | [-0.62, 4.12] | 0.145 | 2.86 | [-0.58, 5.14] | **0.015** |
| FTP SUVR | -9.61 | [-13.18, -6.04] | **< 0.001** |  | -- |  | -7.02 | [-10.78, -3.25] | **<0.001** |
| p-tau217 |  | -- |  | -9.29 | [-13.05, -5.53] | **< 0.001** | -6.12 | [-10.02, -2.26] | **0.002** |
| R<sup>2</sup> | 0.328 |  |  | 0.299 |  |  | 0.402 |  |  |
| AIC | 526 |  |  | 530 |  |  | 518 |  |  |

*Adjusted cortical volume models (A) include all 87 patients, while MMSE models (B) include 85 patients. Sex is dummy coded as 0 for Males and 1 for Females so positive estimates indicate higher volumes/MMSE scores in Females. Other variables are centered, not standardized.*
*R<sup>2</sup> = Coefficient of Determination (higher R<sup>2</sup> is better), AIC = Akaike Information Criterion (lower AIC is better).*

### Association between tau biomarkers at the voxel-level

Whole brain analyses showed that p-tau217 concentration was positively associated with FTP-SUVR throughout the neocortex, with strongest correlations surviving stringent FWE correction in temporo-parietal and dorsolateral prefrontal cortices (Figure 3A).

**Fig 3.**
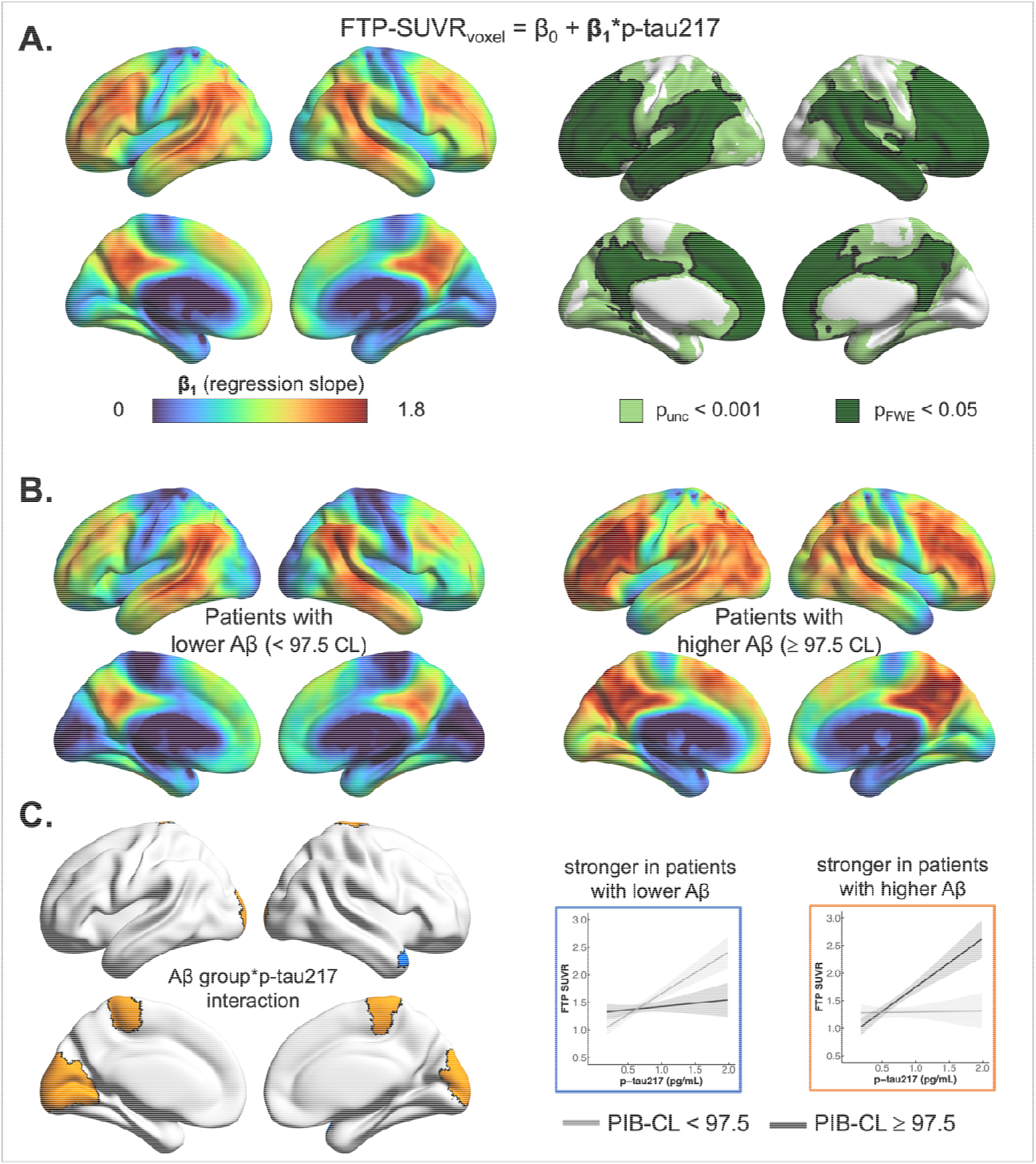
Voxelwise associations between FTP-PET and p-tau217. The association between plasma p-tau217 and FTP-SUVR in each voxel was assessed using a simple regression model in each voxel (FTP-SUVRvoxel = β0 + β1*p-tau217). A: Top-left: non-thresholded maps where β1 values indicate the increase in FTP-SUVR associated with an increase in 1pg/mL of plasma p-tau217. Top-right: thresholded map showing clusters where the plasma-PET association was significant using the two pre-determined threshods (uncorrected p<0.001 and Family-Wise Error corrected p<0.05). B: non-thresholded β_1_ maps estimated in the 2 amyloid groups separately; same colorscale as Panel A. C: interaction results, indicating significant differences between the two maps shown in panel B based on an uncorrected p < 0.001 threshold with a cluster size > 100 voxels. To illustrate these interactions and the direction of the PET-plasma associations, SUVR values were extracted from significant clusters and displayed on scatterplots. Clusters were color-coded to indicate the direction of the interactions. Full 3-dimensional maps, including thresholded and non-thresholded images, are available on neurovault: https://neurovault.org/collections/NLWHVBKP/

Voxelwise interaction analyses indicated that the regional patten of association between p-tau217 and FTP-SUVR was moderated by global amyloid levels (Figure 3B-C) when patients were sub-grouped based on global amyloid Centiloids using a median split (median = 97.5 CLs). In patients with lower amyloid burden [10.4-97.1 CL], p-tau217 was more strongly associated with temporal FTP-SUVR, whereas in patients with higher amyloid [97.5-172.0 CL], p-tau217 was more strongly associated in primary cortices including sensori-motor and visual cortices. Voxelwise interaction models based on other variables (age, sex, APOE-ε4 carriership) did not show significant clusters at the pre-specified significance thresholds.

### Cross-sectional associations between tau biomarkers and downstream markers (brain volume and cognition)

Each single biomarker model showed associations between higher FTP-SUVR or p-tau217 values and lower adjusted cortical volume (p=0.006 and p=0.043, respectively; see full model description in Table 2A) when controlling for age and sex. When both biomarkers were included in the same model, only FTP-SUVR remained significant (p=0.037, versus p=0.34 for p-tau217), and the model did not perform better than the FTP-SUVR only model (ΔR^2^ = +0.009, ΔAIC = +1, see Table 2A).

Separate models showed that higher mean cortical FTP-SUVR and p-tau217 were associated with lower MMSE score (r’s = -0.530 and -0.525, respectively; p’s<0.001, Figure 4A). These associations remained highly significant when controlling for age and sex and when tau biomarkers were included in the same model (model 3 in Table 2A), both remained significant predictors of lower MMSE (p’s<0.002); see full model description in Table 2B. This two-biomarker model showed a substantially increased R^2^ (ΔR^2^ = +0.074) and decreased AIC (ΔAIC = -8), compared to the best single biomarker model (i.e., PET only, see Table 2B).

**Fig 4.**
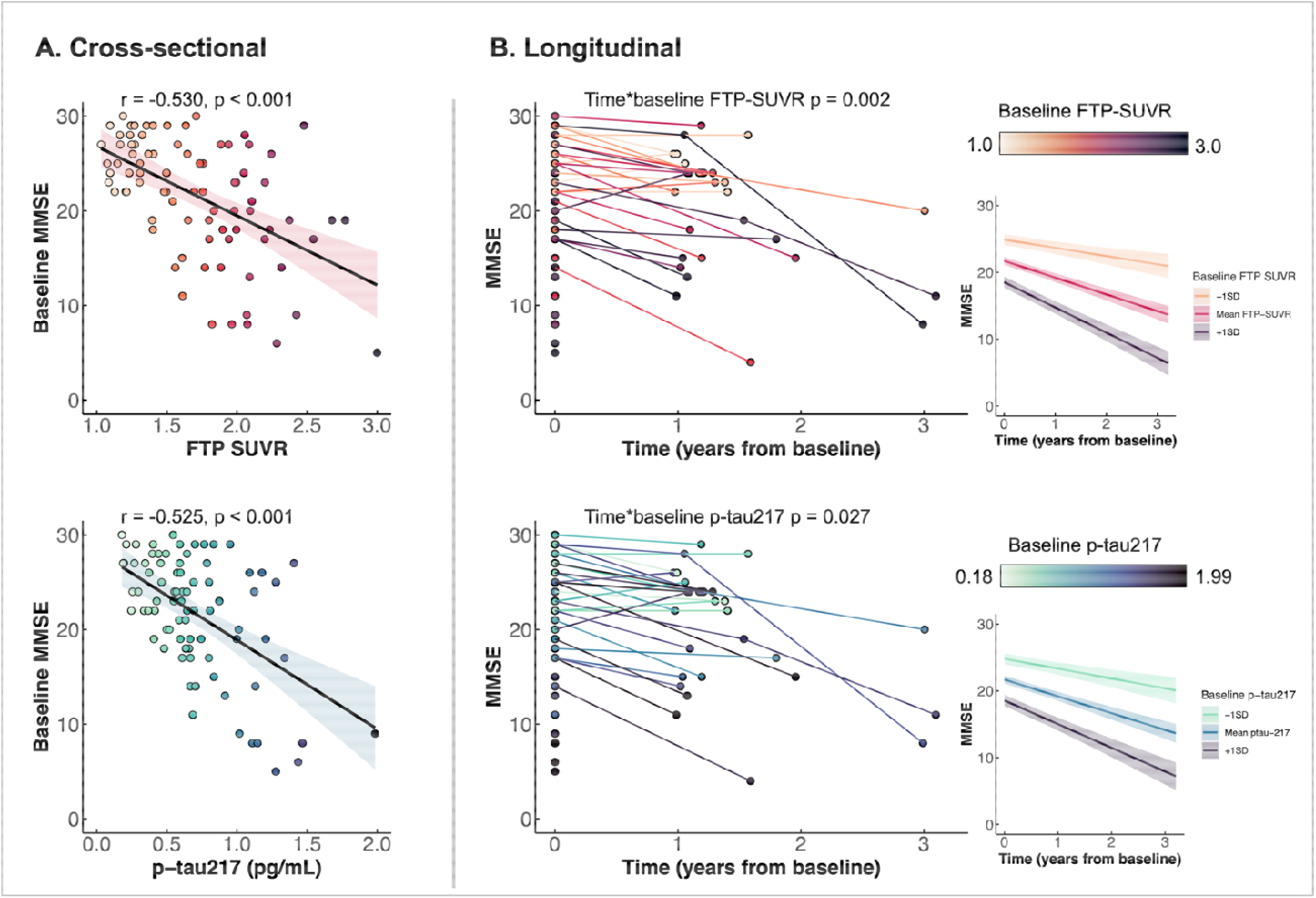
Associations of baseline tau biomarkers with cross-sectional and longitudinal MMSE. Panel B shows raw trajectories (middle) and the output of a linear mixed effect model (right). Number of observations = 85 for cross-sectional analysis (A); n = 116 for longitudinal analysis (B), including all baseline scores + 31 follow up scores from 28 patients

### Associations between baseline tau biomarkers and subsequent MMSE decline

In separate models, baseline FTP-PET and p-tau217 were significantly associated with both baseline MMSE (p’s<0.0001) and change in MMSE over-time (p=0.002 for FTP-PET, p = 0.027 for p-tau217, see full model and coefficients in Figure 4B). However, when both biomarkers were included in the same model, only baseline FTP-PET remained a significant predictor of cognitive decline (p=0.02 versus p=0.36 for p-tau217, Table 3).

**Table 3.** Associations between baseline tau biomarkers and subsequent MMSE decline.

|  | Model 1: PET only |  |  | Model 2: plasma only |  |  | Model 3: both |  |  |
| --- | --- | --- | --- | --- | --- | --- | --- | --- | --- |
| | $\beta$ | 95% CI of $\beta$ | p | $\beta$ | 95% CI of $\beta$ | p | $\beta$ | 95% CI of $\beta$ | p |
| Intercept | 21.7 | [20.6, 22.9] | **< 0.0001** | 21.7 | [20.5, 22.9] | **< 0.0001** | 21.7 | [20.6, 22.8] | **< 0.0001** |
| Time | -2.5 | [-3.30, -1.72] | **< 0.0001** | -2.5 | [-3.4, -1.6] | **< 0.0001** | -2.5 | [-3.3, -1.7] | **< 0.0001** |
| Baseline FTP SUVR | -7.0 | [-9.58, -4.38] | **< 0.0001** |  | -- |  | -4.2 | [-7.4, -1.1] | **0.009** |
| Baseline p-tau217 |  | -- |  | -9.1 | [-12.5, -5.7] | **< 0.0001** | -5.9 | [-9.9, -1.8] | **0.006** |
| Time*Baseline FTP SUVR | -2.8 | [-4.40, -1.19] | **0.002** |  | -- |  | -2.4 | [-4.2, -0.5] | **0.02** |
| Time*Baseline p-tau217 |  | -- |  | -3.0 | [-5.6, -0.5] | **0.027** | -1.3 | [-4.1, 1.5] | 0.36 |
| AIC |  | 703 |  |  | 709 |  |  | 696 |  |
| ICC |  | 0.775 |  |  | 0.735 |  |  | 0.76 |  |
| R<sup>2</sup> (marginal) |  | 0.387 |  |  | 0.349 |  |  | 0.436 |  |
| R<sup>2</sup> (conditional) |  | 0.862 |  |  | 0.828 |  |  | 0.864 |  |

*All linear mixed-effects models included random intercepts for patients. Time is expressed in years and non-centered; FTP and p-tau217 are centered, so model intercepts indicate predicted MMSE score at baseline (time = 0) for average FTP and/or p-tau217 values. Number of patients = 85, number of observations = 116 (31 follow up scores from 28 patients). AIC = Akaike Information Criterion (lower AIC is better); ICC = Intraclass Correlation Coefficient.*

## Discussion

With the recent development of blood-based biomarkers that can detect AD pathology, it is crucial to better understand what these markers reflect and how they compare to more established markers of AD pathophysiology in terms of diagnostic, prognostic, and disease tracking ability. In the present study, we conducted a head-to-head comparison of plasma p-tau217 and tau-PET in amyloid-positive patients with MCI and mild dementia to assess these biomarkers’ relations with each other and with demographic, clinical, and other neuroimaging measures. In our cohort, plasma and PET tau biomarkers were strongly correlated and showed comparable associations with demographic variables and downstream markers (brain volume and cognition), although these relationships tended to be stronger with tau-PET than with plasma p-tau217.

Previous studies have shown that PET and plasma biomarkers may reflect different aspects of tau neuropathology where p-tau217 relates to the presence of soluble phosphorylated tau while tau-PET relates to the aggregation of insoluble helical filaments of tau. This difference would affect the timing of when each biomarker elevates in the AD pathophysiological process and explain why studies have showed that plasma p-tau concentrations elevate before tau-PET signal^11,32^ 11/29/2022 9:50:00 PM. This suggests that plasma p-tau217 should correlate more strongly with markers of upstream pathophysiological events (e.g., amyloid deposition) than with downstream markers (e.g., brain atrophy and clinical worsening). Surprisingly, we did not observe this pattern; instead, tau-PET tended to be more strongly related to both upstream and downstream measures, compared to plasma p-tau217. This pattern suggests that, rather than a difference in the timing of these biomarkers, they might defer in terms of signal-to-noise ratio. This hypothesis is consistent with recent observations that plasma p-tau217 concentrations are influenced by non-neurodegenerative comorbidities, including renal and hepatic diseases.^33–35^ These factors could result in plasma-specific measurement error that would weaken associations with other variables.

Per study design, all the patients included in this study were visually amyloid-PET positive; yet, a wide range of amyloid-PET levels was observed, as evidenced by Centiloid values ranging from 10 to 172 (median = 97.5, close to the 100 CL anchor point defined as the average value observed in patients with mild AD dementia^36^). However, this major variability in Centiloids was very weakly associated with tau markers: r=0.26 for PET (p=0.02), r=0.10 for plasma (p=0.36). This finding is not in line with the common idea that plasma p-tau biomarkers are more closely associated with early markers of AD such as amyloid^1,6,9^, while tau-PET is more closely associated with downstream markers of disease progression such as cognitive decline and brain atrophy^37^, as discussed in the previous paragraph. Consistent with our data, an independent study of amyloid-positive patients found that p-tau217-to-tau-PET association was stronger than p-tau217-to-amyloid-PET^38^. Furthermore, it has been shown that associations between plasma p-tau217 and PET markers evolve over the course of the disease^5^ : plasma p-tau217 is associated with amyloid-PET in the earliest stages of AD, when tau burden is restricted to the medial temporal lobe. In later stages, plasma p-tau217 is related to both amyloid-PET and tau-PET, although more strongly associated with tau-PET. Therefore, our finding of a moderate (r=0.61) association between plasma p-tau217 and tau-PET, but not between plasma p-tau217 and amyloid-PET is likely due to the lack of cognitively unimpaired and amyloid-negative impaired patients in our cohort.

Voxelwise analyses showed that plasma p-tau217 was strongly correlated with tau-PET signal in a typical AD pattern encompassing temporo-parietal and dorsolateral prefrontal cortices. Interestingly, the regional pattern of association differed in individuals with lower vs. higher amyloid levels. In patients with moderate amyloid burden (lower half of Centiloid distribution), plasma p-tau217 was more strongly associated with tau-PET in the temporal lobes, whereas plasma p-tau217 was more strongly related to tau-PET signal in sensorimotor and visual cortices in patients with elevated amyloid burden.

While our modest sample size results in relatively low power to detect interactions, it is interesting to note that these regions (Figure 3B) are particularly relevant as they mirror the progression of tau pathology throughout the brain, from medial temporal regions (early Braak stage regions) to primary cortices (in Braak Stage VI).^39,40^

Our results are congruent with existing evidence linking tau biomarkers with downstream measures of AD pathophysiology. ^22–24^ In single biomarker analyses, tau-PET and plasma p-tau217 were associated with lower cortical volume and MMSE. Yet, models that included both tau biomarkers differed: while both markers independently contributed to cross-sectional MMSE scores, plasma p-tau217 did not significantly account for brain volume or longitudinal MMSE decline once tau-PET was in the model. Taken together, these findings suggest that, while tau-PET tends to be more robustly associated with downstream neurodegeneration and cognitive decline, tau-PET and plasma p-tau217 seem to reflect closely associated pathophysiological processes and could both be helpful to estimate disease stage and provide prognostic information.

Previous studies have shown that, because amyloid-PET signal typically increases at a consistent rate across individuals, amyloid-PET levels reflect the duration of amyloid positivity (or amyloid ‘chronicity’)^41,42^, an index of how long patients have been on the pathophysiological pathway. In the present study, our data suggests that at the earlier stages of the AD cascade (i.e., in patients with moderately positive Centiloid values), plasma p-tau217 reflects earlier stages of tau spread, when tau is still mainly limited to the temporal lobes. In later stages (i.e., in patients with highly positive Centiloids), plasma p-tau217 then tracks tau spread to later regions. In summary our findings support the hypothesis that plasma p-tau217 reflects early as well as late stages of tau spread.

Overall, the relationships we observed between tau-PET and demographic and clinical variables are consistent with previous reports in amyloid-positive patients. In line with previous PET and neuropathology studies,^43,44^ both younger age of onset and female sex were associated with higher tau-PET burden. In contrast, mean cortical tau-PET SUVR was independent of APOE-ε4 carriership, consistent with converging evidence that APOE-ε4 has a focal effect on tau accumulation in the medial temporal lobe, rather than an impact on global tau burden.^17,45,46^ Plasma p-tau217 showed similar pattern of associations with age, sex, and APOE-ε4, although the correlation with age was weaker with plasma p-tau217 (r=-0.44) than with tau-PET (r=-0.68, difference: p=0.003). Altogether, these results suggest that levels of plasma p-tau217 and tau-PET are driven by similar factors in our cohort.

Our study has limitations. Because our sample size is relatively small, especially for longitudinal analysis, we may have limited power to detect significant associations between plasma p-tau217 and other measures. Our sample mainly consisted of a single cohort of highly selected patients recruited in an academic setting and lacked racial and ethnic diversity (95% White). Additional studies in larger and more diverse cohorts are needed assess and validate these findings for the use plasma p-tau217 as a scalable biomarker.

With the emerging evidence suggesting the value of plasma p-tau217 and the difference in terms of cost, invasiveness, and accessibility, it is inevitable to question whether plasma p-tau217 could replace tau-PET as a measure of tau pathology for diagnostic and prognostic purposes or in the context of clinical trials. While p-tau217 concentrations provided some information on downstream disease processes, tau-PET could be valuable in providing information on underlying disease processes beyond the global cortical measure that was used in this study for the sake of PET-plasma comparison. For a given value of p-tau217 concentration, we observed great variability in the global tau burden and regional pattern of tau-PET signal (Figure 1). Previous studies have shown that the regional patterns of tau-PET signal not only closely mirror current the clinical representation of AD related clinical syndromes ^47^ but also with future patterns of brain atrophy^23,24^ as well as are correlated with domain-specific cognitive impairments^47–49^. This suggests that p-tau217 could serve as an important biomarker to assess the presence of tau pathology and disease severity, but tau-PET is advantageous for tracking regional specific tau pathology.

In conclusion, in this direct comparison of plasma and PET tau biomarkers, we show that both tau biomarkers have similar patterns of associations with demographic and clinical variables and with downstream markers of disease progression, although associations tended to be stronger with PET than plasma. These findings suggest that, beyond assessing the presence of AD, plasma biomarkers could also inform on disease severity; yet broader assessment and validation are required for more extensive use of blood-based biomarkers.

## Acknowledgements

We thank all patients, caregivers, and families for their commitment. We thank Amelia Strom, Daniel Schonhaut, and Sarah Ackley for their help during data analyses and manuscript preparation. Avid Radiopharmaceuticals enables the use of Flortaucipir tau-PET tracer but did not provide direct funding and were not involved in the data analysis or interpretation. Plasma p-tau assay results were donated by Lilly Research laboratories.

## Appendix 1

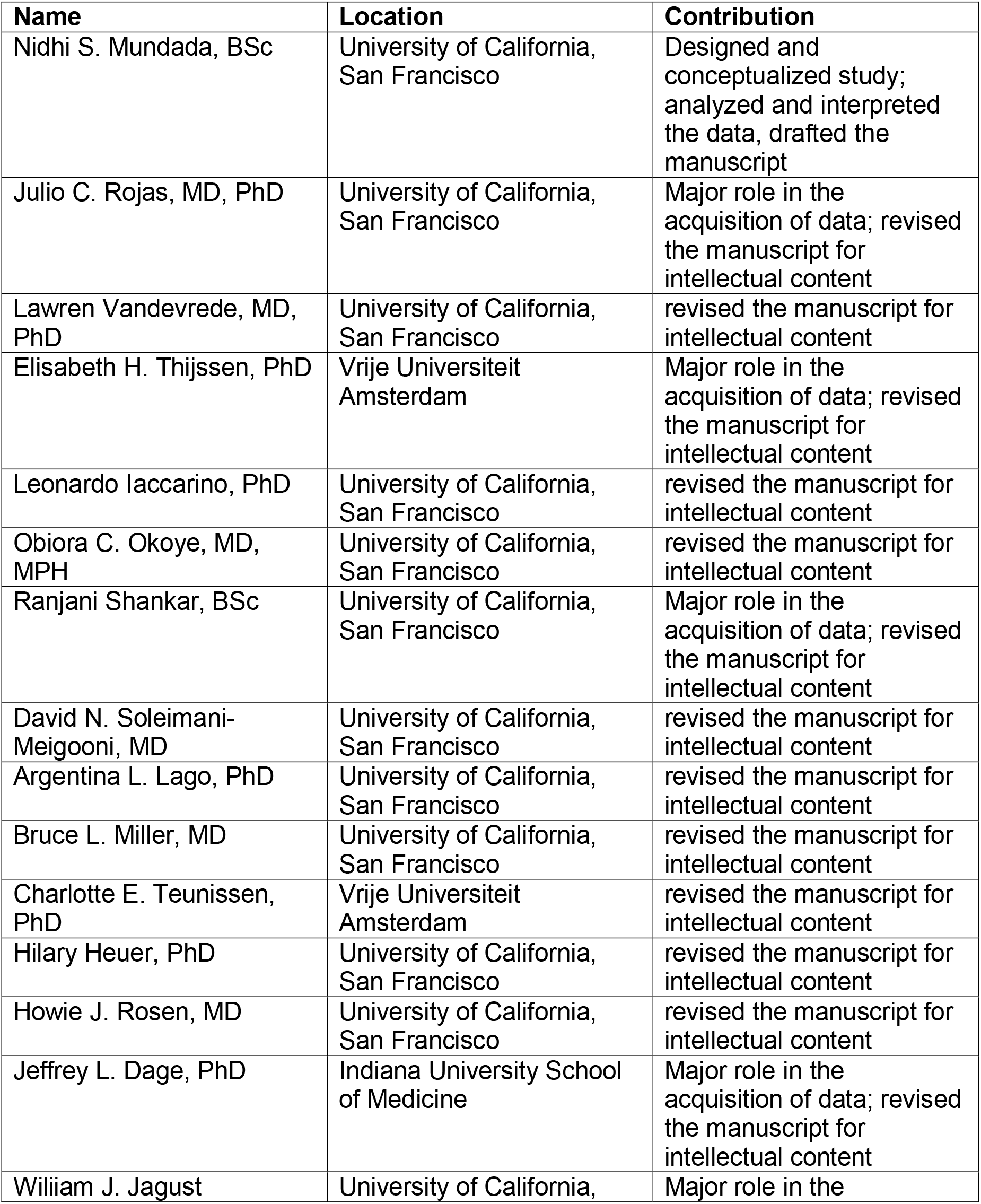
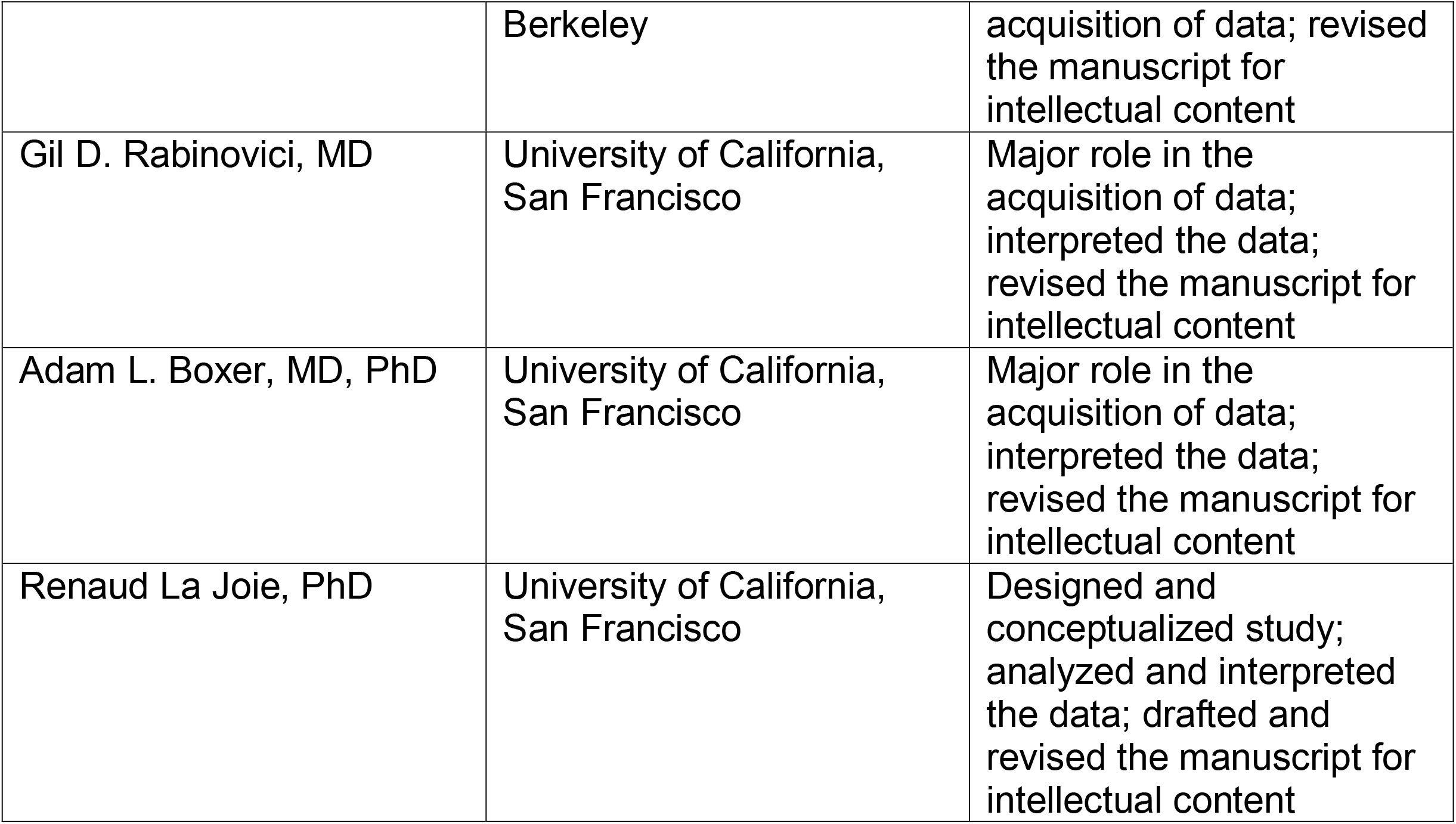

