## Supplementary for "Head-to-head comparison between plasma p-tau217 and Flortaucipir-PET in amyloid-positive patients with cognitive impairment"

### Supplementary Materials

#### Supplementary Fig 1. Study flow-chart

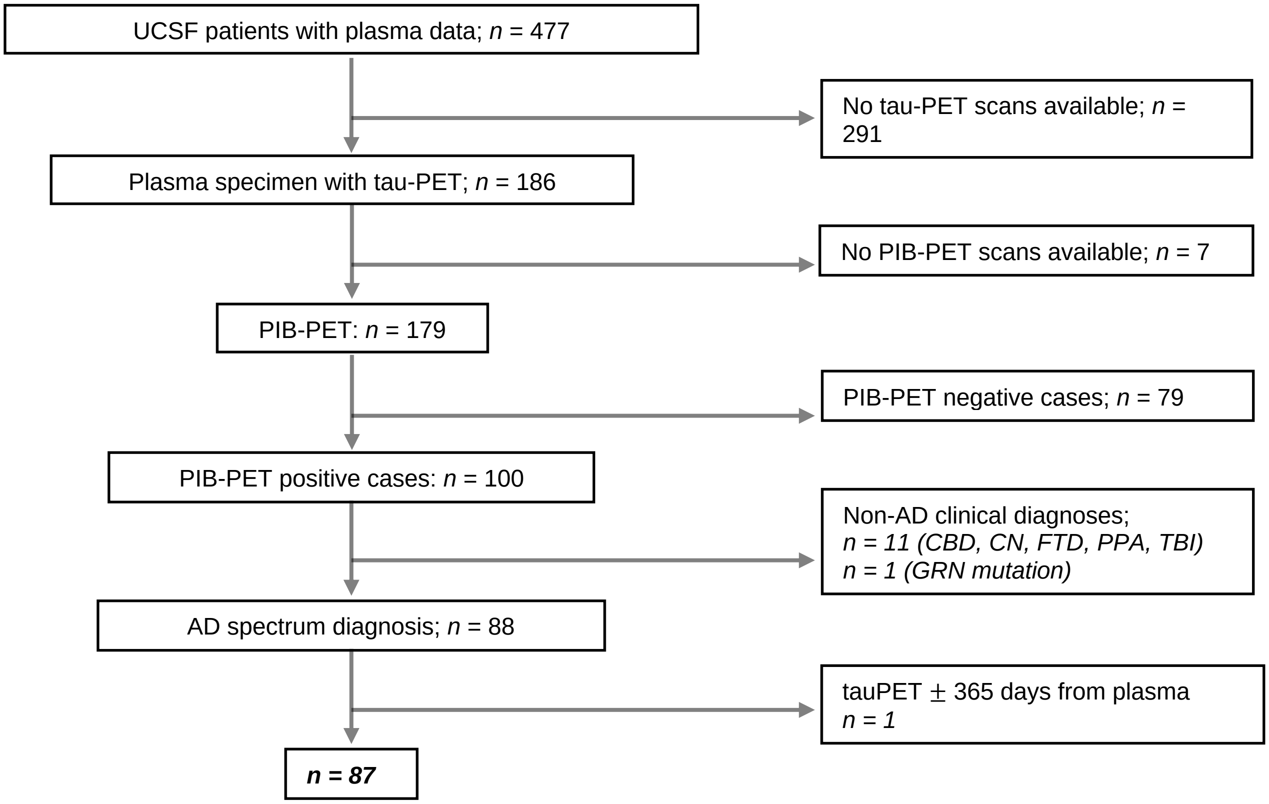


#### Supplementary Table 1. Comparison of the associations of plasma p-tau217 and FTP-PET with other variables using bootstrapping.

| | **Age** | | **Sex** | | **APOE (number of e4 alleles)** | | **Amyloid-PET (CLs)** | |
|---|---|---|---|---|---|---|---|---|
| | r | p | d | p | $\eta^2$ | p | r | p |
| FTP-PET | -0.68 | <0.001 | 0.78 | <0.001 | 0.002 | 0.91 | 0.26 | 0.02 |
| p-tau217 | -0.44 | <0.001 | 0.53 | <0.001 | 0.015 | 0.52 | 0.10 | 0.36 |
| | $\Delta$r [95%CI] | p | $\Delta$d [95%CI] | p | $\Delta\eta^2$ [95%CI] | p | $\Delta$r [95%CI] | p |
| Bootstrap | -0.24 [-0.40, -0.08] | 0.003 | 0.25 [-0.16, 0.72] | 0.21 | -0.013 [-0.08, 0.06] | 0.81 | 0.16 [-0.01, 0.34] | 0.09 |

#### Supplementary Table 2. Associations of plasma p-tau217 and FTP-PET with demographics and other variables after log-transformation or based on rank-based statistics.

| Variables | | Raw | | Log-transformed | | Non-parametric | |
|---|---|---|---|---|---|---|---|
| FTP – p-tau217 | | r | 0.61 | r | 0.66 | $\rho$ | 0.64 |
| | | p | <.001 | p | <.001 | p | <.001 |
| Age | FTP | r | -0.68 | r | -0.67 | $\rho$ | -0.68 |
| | | p | <.001 | p | <.001 | p | <.001 |
| | p-tau217 | r | -0.44 | r | -0.46 | $\rho$ | -0.48 |
| | | p | <.001 | p | <.001 | p | <.001 |
| Sex | FTP | d | 0.78 | d | 0.80 | rrb | 0.39 |
| | | p | <.001 | p | <.001 | p | <.001 |
| | p-tau217 | d | 0.53 | d | 0.56 | rrb | 0.29 |
| | | p | 0.016 | p | 0.01 | p | 0.02 |
| APOE4 | FTP | $\eta^2$ | 0.002 | $\eta^2$ | 0.001 | $\varepsilon^2$ | 0.002 |
| | | p | 0.91 | p | 0.95 | p | 0.92 |
| | p-tau217 | $\eta^2$ | 0.015 | $\eta^2$ | 0.007 | $\varepsilon^2$ | 0.003 |
| | | p | 0.52 | p | 0.74 | p | 0.87 |
| PIB CLs | FTP | r | 0.26 | r | 0.26 | $\rho$ | 0.19 |
| | | p | 0.02 | p | 0.015 | p | 0.08 |
| | p-tau217 | r | 0.10 | r | 0.21 | $\rho$ | 0.07 |
| | | p | 0.36 | p | 0.057 | p | 0.52 |
| MMSE | FTP | r | -0.530 | r | -0.530 | $\rho$ | -0.509 |
| | | p | <.001 | p | <.001 | p | <.001 |
| | p-tau217 | r | -0.525 | r | -0.507 | $\rho$ | -0.443 |
| | | p | <.001 | p | <.001 | p | <.001 |

*For log-transformed associations, only FTP-SUVR and p-tau217 concentrations were log-transformed; other all values remained untransformed. For non-parametric measures, spearman's test was used for continuous variables (ρ = Spearman's rho (rank-based)) coefficient), Mann-Whitney U test for binary variables (rrb = rank biserial r), Kruskal-Wallis ANOVA for categorical variables with more than 2 categories ($\varepsilon^2$: effect size from Kruskal-Wallis non-parametric ANOVA)*

#### Supplementary Methods: R code

**Code for bootstrapped resampling**
Example R code below showing difference in corresponding effect sizes between tau-PET and age and p-tau217 and age:

```
library(cocor)
library(boot)
library(boot.pval)

data_age <- data.frame(age=bldf$bl_age, pet=bldf$bl_FTP,
plasma=bldf$bl_pTau)

boot.function <- function(data_age,indices
  data_age <- data_age[indices,]
  corr1 <- cor.test(data_age$age,data_age$pet)
  corr2 <- cor.test(data_age$age,data_age$plasma)
  diff <- corr1$estimate-corr2$estimate
  return(diff)
}

set.seed(12345)
boot.out <- boot(data_age,boot.function,R=1000)

# CI
boot.ci(boot.out,type="perc")
# P-value
boot.pval(boot.out,type="perc")
```

The above code was repeated similarly for amyloid-PET Centiloids; however, the difference in effect sizes was calculated cohen’s d for sex and eta-squared for APOE-ε4.

**Code for linear mixed effect models**

```
library(lme4)
library(lmerTest)
library(nlme)
library(dplyr)

df <- read.table("05-Longitudinal_MMSE_NM_06-25.csv", header =
TRUE, sep = ",")
mmsedf_lme <- df[c(1,4,7,8)]
colnames(mmsedf_lme) <-
c("ID","time_from_first_mmse","bl_FTP","bl_pTau")

### Mean Center function
len <- length(colnames(mmsedf_lme))

center_scale <- function(x) {
  scale(x, scale = FALSE)
}

# Select columns that don't need centering
mmsedf_lme_nocentering <- mmsedf_lme[,1]

# Select columns that need centering
mmsedf_lme_centering <-
center_scale(select_if(mmsedf_lme[,3:len], is.numeric))

# Bind non-mean centered data and mean centered data into one
dataset
mmsedf_lme_centered <- cbind(mmsedf_lme_nocentering,
mmsedf_lme_centering)

# Model 1: LME model with FTP
longitmmse_FTP <- lmer(MMSE ~ 1 + time_from_first + bl_FTP +
time_from_first*bl_FTP + (1 | ID), data = mmsedf_lme_centered)
summary(longitmmse_FTP)

# Model 2: LME model with p-tau217
longitmmse_pTau <- lmer(MMSE ~ 1 + time_from_first + bl_pTau +
time_from_first*bl_pTau + (1 | ID), data = mmsedf_lme_centered)
summary(longitmmse_pTau)

# Model 3: LME model with FTP and p-tau217
longitmmse_pTauFTP <- lmer(MMSE ~ 1 + time_from_first + bl_pTau
+bl_FTP + time_from_first*bl_pTau + time_from_first*bl_FTP + (1
| ID), data = mmsedf_lme_centered)
summary(longitmmse_pTauFTP)
```
